# Trends in cigarette and e-cigarette use among youth: Findings from the 2014-2023 China Youth Tobacco Survey in Heilongjiang Province

**DOI:** 10.1101/2025.09.12.25335638

**Authors:** Linlin Jiang, Ying Wang, Xinbo Di

## Abstract

**Introduction:** To identify intervention priorities for reducing tobacco use among youth in Heilongjiang, this study examined provincial epidemiological patterns of cigarette and e-cigarette adoption from 2014 to 2023, providing evidence-based foundations for timely provincial school tobacco control strategies.

**Methods:** School-based cross-sectional surveys were conducted in 2014, 2019, 2021 and 2023. A randomized multistage stratified cluster sampling design was employed to obtain representative samples of middle school students (MSs) and high school students (HSs).

**Results:** In 2019, 2021 and 2023, the current cigarette and e-cigarette use rates among MSs and HSs were 6.79%, 5.45%, and 4.70%, and 3.61%, 5.46%, and 2.07%, respectively. The current dual-use rates for cigarettes and e-cigarettes were 1.74%, 2.83%, and 1.03%, respectively. The current rates for either cigarette or e-cigarette use were 8.70%, 8.10%, and 5.76%, respectively. Among current cigarette users of MSs and HSs, the rates of e-cigarette use first were 15.59%, 12.75%, and 21.80%, respectively. Among cigarette and e-cigarette dual-users of MSs and HSs, the rates of e-cigarette use first were 17.46%, 14.99%, and 28.41%, respectively.

**Conclusions:** From 2014 to 2023, based on results from four province-wide surveys, cigarette use rates among MSs and HSs in Heilongjiang declined, while e-cigarette use rates showed a peak in 2021 before decreasing. Policies such as “smoke-free schools” and e-cigarette regulation were beginning to show results. Joint efforts between families and schools, as well as collaboration among multiple departments were essential to ensure effective implementation of tobacco control measures and would be the focus of future tobacco control work.

## Introduction

The World Health Organization (WHO) has identified tobacco use as the largest current public health threat globally in its latest report on the global tobacco epidemic, making tobacco control a priority for global public health^1^. The 2018 Chinese Adult Tobacco Survey revealed that 26.6% of individuals aged ≥15 years in China were smokers ^2^, suggesting that over 300 million people smoke tobacco ^3^. In 2019 alone, tobacco-attributed deaths in China reached 2.7 million ^4^. Additionally, the substantial disease burden and economic losses caused by secondhand smoke exposure cannot be overlooked ^5^. Extensive evidence has established a positive correlation between smoking duration and tobacco-related harm: earlier probability of successful cessation, thereby intensifying tobacco’s harmful effects on the body ^6^. Research on youth tobacco use trends, considered the ‘critical predictor’ of adult tobacco epidemics, is therefore particularly important. Reducing new smoker emergence and decreasing existing smoker numbers are recognized as effective strategies for lowering smoking rates, with youth-focused interventions offering the greatest potential for preventing new smokers ^7^. Focusing on youth tobacco use enables earlier intervention in adult tobacco control efforts, making youth smoking prevention a crucial factor in reducing population smoking rates and smoking-related disease incidence ^8^.

Since the WHO Framework Convention on Tobacco Control (FCTC) took effect in China, tobacco control initiatives have received heightened attention from governmental bodies across all levels and diverse sectors. The ‘Healthy China 2030’ blueprint explicitly advocates to ‘comprehensively advance tobacco control convention compliance and intensify tobacco control efforts’ ^9^. Consequently, tobacco control—particularly youth tobacco control—has demonstrated steady progress. Recent national youth tobacco surveillance data indicate a decline in current cigarette use rates among Chinese middle and high school students, from 5.9% in 2019 to 4.2% in 2023, reflecting a consistent downward trajectory ^10–12^. While youth tobacco control shows positive development, concerns regarding youth e-cigarette usage have emerged within tobacco control discourse. According to the 2019 China Youth Tobacco Survey, the current e-cigarette use rate among Chinese MSs and HSs stood at 2.8% ^10^, rising to 3.6% in 2021 ^11^, before declining to 2.4% in 2023 ^12^. As novel tobacco products, e-cigarettes have gained global popularity through innovative designs and diverse flavor profiles ^13–14^. Since their introduction in China in 2004 ^15^, these products have garnered significant interest among youth populations. However, the WHO maintains that e-cigarettes pose public health risks, are not smoking cessation aids, and require stringent regulation to prevent harm to non-smokers and youth populations ^16^.

Tobacco control researchers worldwide have documented strong associations between e-cigarette and cigarette use. Existing studies demonstrate that e-cigarette use correlates with both smoking intention and subsequent smoking behavior ^17–18^. Furthermore, the use of electronic nicotine delivery systems (ENDS) increases relapse risk among former smokers ^19^. A national tobacco survey targeting Chinese middle and high school students revealed that e-cigarette use among never-smokers and ever-smokers positively correlates with susceptibility to cigarette use. This study further established a dose-response relationship between frequency of e-cigarette use and susceptibility to cigarette use ^20^.

Regarding youth tobacco use epidemiology, although tobacco prevalence surveys have been conducted across different years in Heilongjiang Province, a significant research gap persists in provincial-level analyses of youth tobacco trends—specifically in either cigarette or e-cigarette use and dual-use patterns. Furthermore, as a high-latitude region characterized by prolonged harsh winters, the distinct geographical and climatic conditions have shaped unique lifestyle patterns in the Heilongjiang area. Region-specific tobacco control strategies for youth need to be developed to facilitate provincial intervention initiatives. This study investigates decadal trends (2014-2023) in cigarette and e-cigarette use among MSs and HSs in Heilongjiang Province. It aims to consolidate achievements from smoke-free schools, health-promoting schools, and smoke-free households, address critical data deficiencies, and evaluate stage-specific outcomes of these initiatives while providing evidence-based support for updating and implementing effective provincial youth tobacco control policies.

## Methods

### Study Design and Participants

periods: 2014, 2019, 2021, and 2023. The survey encompassed middle schools (Grades 7-9) and high schools (Grades 10-12), with high schools including both academic high schools (AHS) and vocational high schools (VHS). We employed a cross-sectional design with stratified multistage cluster random sampling, stratifying the entire province into urban and rural domains. Drawing on data from the Chinese Sixth and Seventh National Population Census, we comprehensively considered factors including urban-rural population distribution and minimum required sample sizes. Using probability proportional to size (PPS) sampling methodology, we selected 10 surveillance counties/districts annually (five urban areas, five rural areas). Within each selected district or county, we employed PPS methods to select three middle schools and three high schools (including two academic high schools and one vocational high school) from all local public and private schools. (Note: Only middle schools were sampled in 2014). Subsequently, one class from each grade was randomly selected within sampled schools, with all students in the selected class serving as potential participants. The study obtained informed consent and received approval from the Ethics Committee of the Chinese Center for Disease Control and Prevention (China CDC). All information regarding students and schools remained strictly confidential throughout the study period.

### Procedures and definitions

The survey employed an on-site investigator model where students present in sampled classrooms completed anonymous paper questionnaires under supervision, with teachers and school personnel absent to ensure unimpeded and truthful responses. Additionally, we implemented a comprehensive tripartite quality control framework spanning preliminary, ongoing, and final phases. This framework operated through tiered training prior to fieldwork, real-time quality monitoring during data collection, and post-survey verification that included questionnaire auditing, provincial-level validation, and cross-verification during data cleaning. Systematic correction of missing values, outliers, and logical inconsistencies ensured data authenticity, completeness, and end-to-end traceability. The instrument adapted the Global Youth Tobacco Survey (GYTS) core module while incorporating China-specific items to comprehensively assess tobacco use behaviors and critical control indicators.

In this study, current cigarette smokers were defined as students reporting cigarette use on at least one day within the preceding 30 days, as assessed by the question: “How many days have you smoked cigarettes in the past 30 days?” Response options included: zero, one to two, three to five, six to nine, 10 to 19, 20 to 29, and all 30 days. Students selecting more than zero days were classified as current smokers. Similarly, current e-cigarette users were identified through the question: “How many days have you used e-cigarettes in the past 30 days?” with identical response categories. Students selecting more than zero days were defined as current e-cigarette users. Weekly pocket money was assessed via the question: “On average, how much money do you have available for personal spending (regardless of how you spend it) each week?” Response options included: no pocket money; ≤10 RMB; 11-30 RMB; 31-50 RMB; and >50 RMB. Parental smoking status was determined by the question: “Do either of your parents smoke?” with response options: neither smokes; one parent smokes; both parents smoke. Teacher smoking status was defined by the question: “During the past 30 days on school premises (both indoors and outdoors), how frequently did you observe teachers smoking?” Response options included: almost daily; sometimes; never; and unknown. General smoking observations on campus were assessed by the question: “In the past 30 days, did you witness anyone smoking inside school buildings or anywhere on school grounds?” with response options: Yes or No. Peer smoking influence was captured through: “How many of your closest friends smoke?” Response options included: none; some; most; and all. Perceived harm of secondhand smoke was evaluated through: “Do you believe exposure to others’ cigarette smoke harms your health?” Response options included: definitely not; probably not; probably; and definitely. Students who chose “definitely not” or “probably not” were classified as lacking awareness of secondhand smoke harms, while those who chose “probably” or “definitely” were classified as having accurate awareness of secondhand smoke harms.

### Data analysis

Given the complex multistage stratified cluster random sampling design, we implemented comprehensive weighting procedures for all analyses. We performed data cleaning, sample weighting, parameter estimation, and standard error calculations using SAS 9.4 software. Categorical variables are presented as frequencies with weighted proportions (%). We calculated the final weight for each participant by multiplying the sample selection weight, non-response adjustment coefficient, and post-stratification factors. Post-stratification benchmarks were derived from official enrollment statistics provided by the Heilongjiang Provincial Department of Education for the survey year, stratified by school type, grade level, and gender. All analyses incorporated these final weights to produce weighted parameter estimates. We conducted group comparisons using the Rao-Scott χ² test, which accounts for complex sampling design, with statistical significance defined as *P* < 0.05. We examined factors associated with exclusive cigarette or e-cigarette use among youth using both univariate and multivariate logistic regression models. All statistical tests were two-tailed with a significance level of α = 0.05.

## Results

### Demographic characteristics

In 2014, a total of 4,152 valid questionnaires were collected, with 2,099 males (51.60%) and 2,053 females (48.40%), and 1,319 (35.81%) from urban areas and 2,833 (64.19%) from rural areas. Grade-level distribution included 1,429 grade seven students (32.6%), 1,361 grade eight students (33.28%), and 1,362 grade nine students (34.11%). In 2019, 2021 and 2023, a total of 8,091, 8,329 and 7,252 valid questionnaires were collected separately. With 4,159 males (51.64%) and 3,932 females (48.36%) in 2019, and 4,159 males (51.64%) and 3,932 females (48.36%) in 2021, as well as 3,789 males (51.38%) and 3,463 females (48.62%) in 2023. With 3,923 (44.33%), 4,023 (61.03%) and 3,656 (69.15%) from urban areas and 4,168 (55.67%), 4,306 (38.97%) and 3,596 (30.85%) from rural areas each year. Among MS students (n=4,100, 4,153, 3,735), grade-level distribution comprised 1,379, 1,427 and 1,306 grade seven (33.00%, 39.48% and 44.35%), and 1,330, 1,363, 1,261 grade eight (33.52%, 43.27%, 28.08%), and 1,391, 1,363, 1,168 grade nine students (33.47%, 17.25%, 27.57%). Among HS students (n=3,991, 4,176, 3,517), there were 1,406, 1,448, 1,195 grade 10 (30.9%, 35.43%, 33.49%), and 1,388, 1,381, 1,156 grade 11 (34.76%, 34.36%, 33.18%), and 1,197, 1,347, 1,166 grade 12 students (34.34%, 30.21%, 33.32%). (Table 1)

**Table 1.**
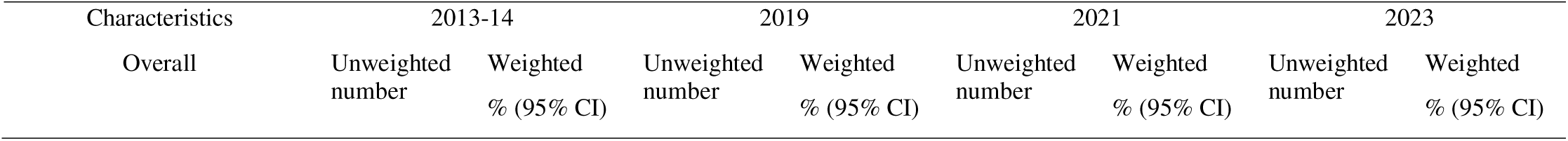
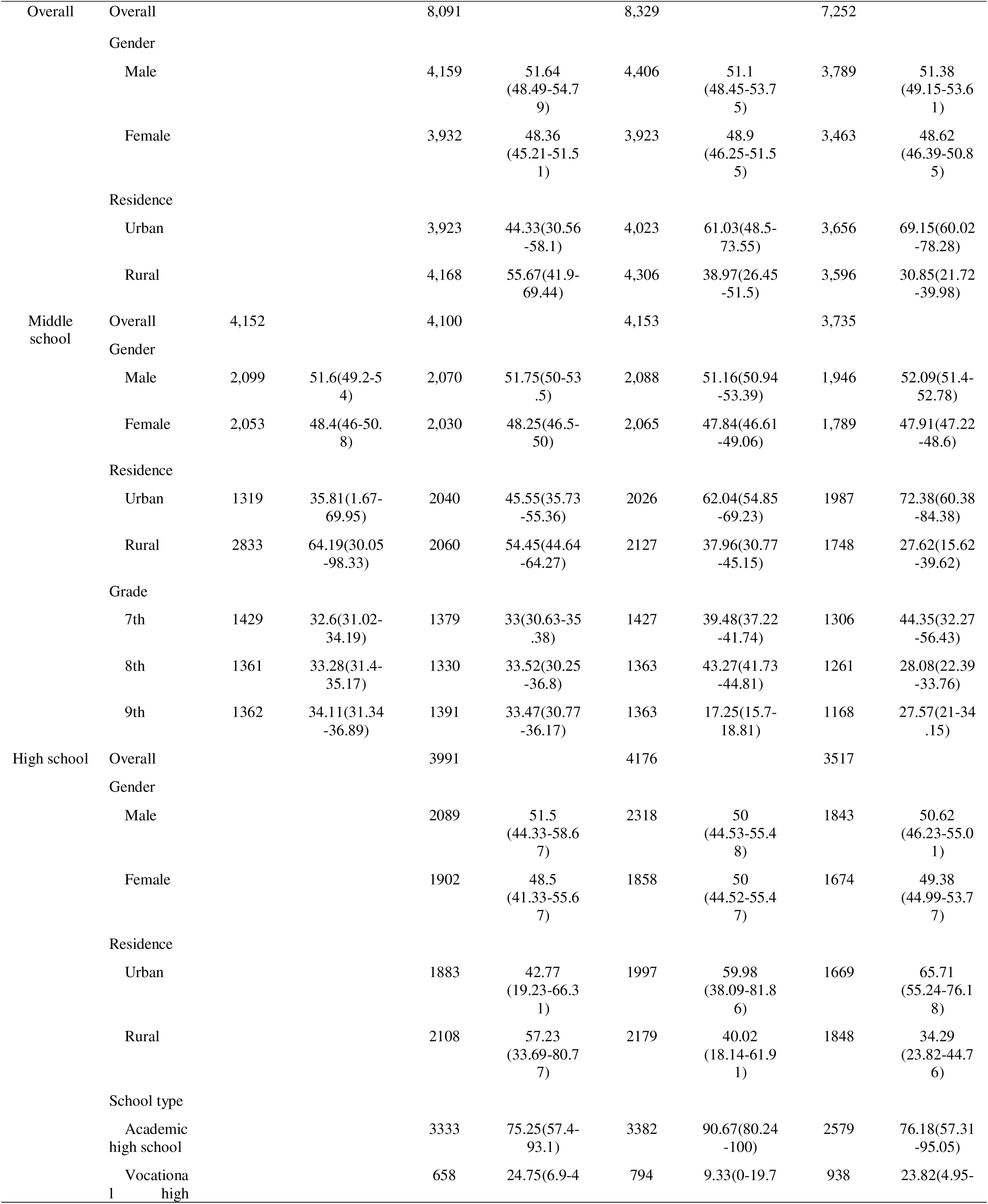
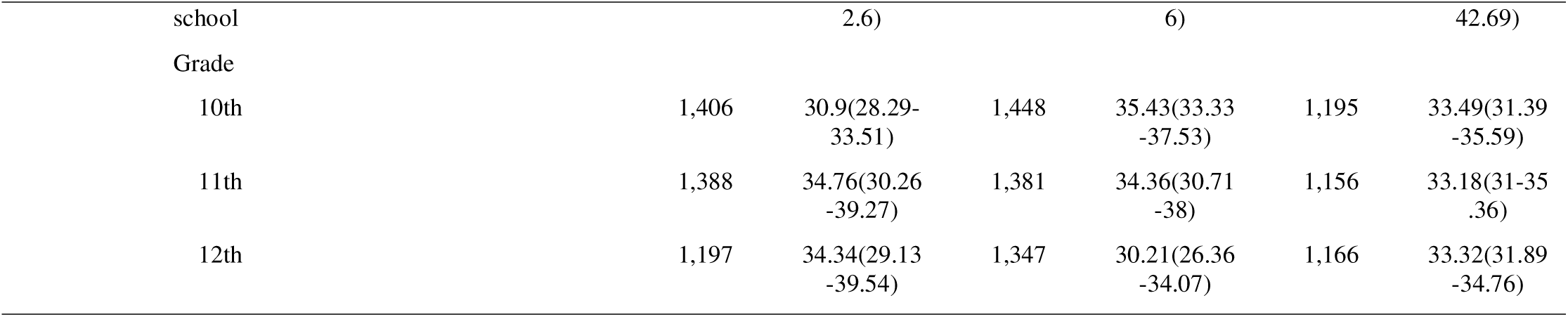
Sociodemographic characteristics of middle school and high school students in Heilongjiang, China.

### Cigarette use among youth in Heilongjiang Province during 2014-2023

The current cigarette use rate among MS and HS students in Heilongjiang Province demonstrated a declining trend from 2019-2023, decreasing from 6.79% (95% *CI*: 4.00%-9.58%) in 2019 to 5.45% (95% *CI*: 3.19%-7.72%) in 2021, and further to 4.70% (95% *CI*: 2.67%-6.73%) in 2023. Among MS students specifically, the prevalence declined consistently from 4.22% (95% *CI*: 1.59%-6.85%) in 2014 to 3.38% (95% *CI*: 1.44%-5.32%) in 2019, 2.05% (95% *CI*: 0.82%-3.29%) in 2021, and 1.60% (95% *CI*: 0.79%-2.41%) in 2023. For HS students, the rates were 11.19% (95% *CI*: 6.92%-15.45%) in 2019, 8.98% (95% *CI*: 5.97%-11.99%) in 2021, and 8.02% (95% *CI*: 5.38%-10.67%) in 2023. (Table 2)

**Table 2.**
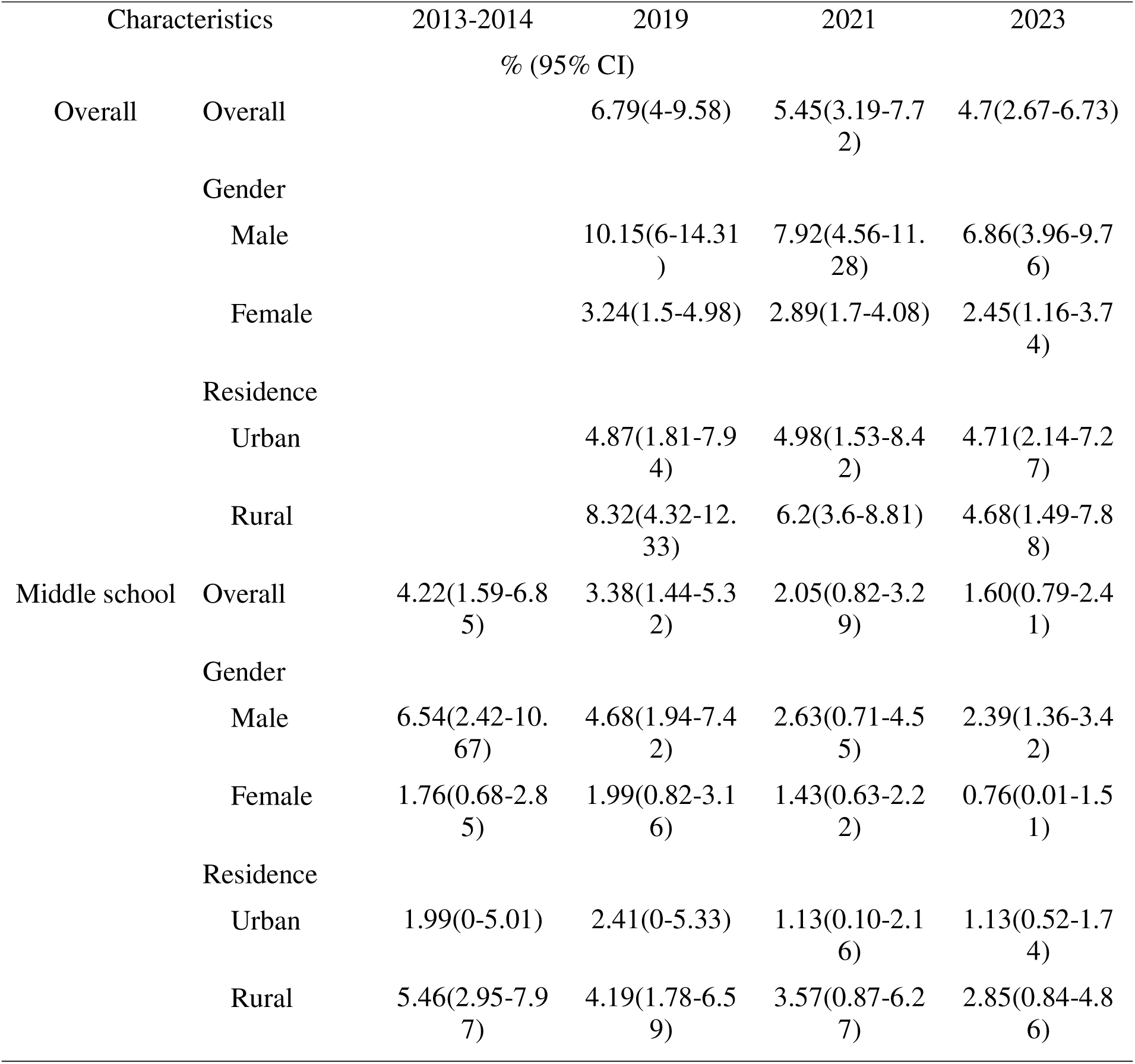
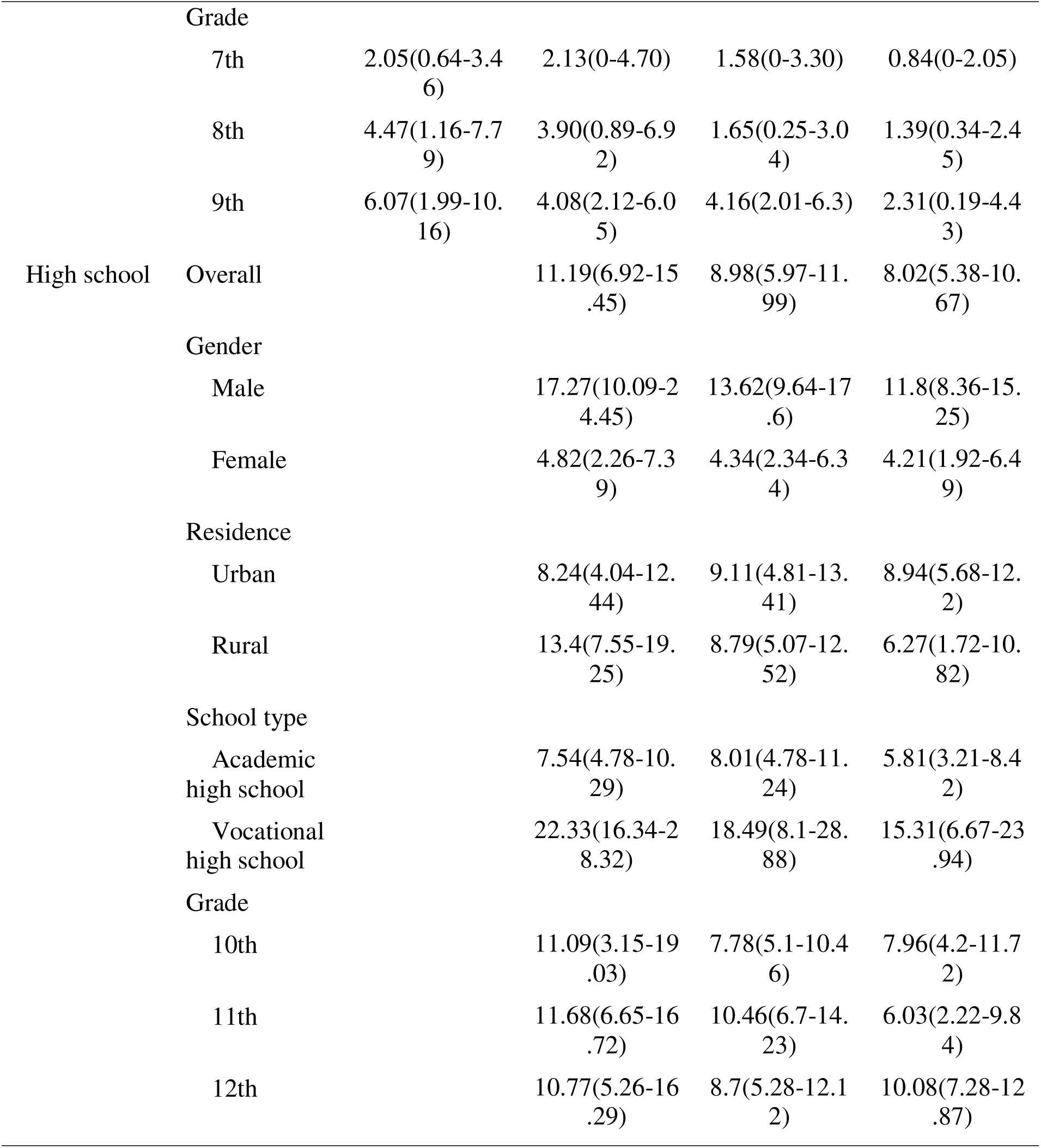
Current use of cigarette among MS students and HS students in Heilongjiang, China.

### E-cigarette use among youth in Heilongjiang Province during 2014-2023

The current e-cigarette use rate of MS and HS students in Heilongjiang Province during 2019-2023 was 3.61%(95%*CI*: 2.58%-4.65%)、5.46%(95%*CI*: 3.33%-7.58%)、2.07%(95%*CI*: 1.23%-2.92%), respectively. Among MS students, the rate was 1.32%(95% *CI*: 1.08%-1.55%)、2.65%(95% *CI*: 1.05%-4.26%)、2.67%(95% *CI*: 1.3%-4.04%) and 1.6%(95% *CI*: 1.15%-2.05%) from 2014 to 2023, respectively. Among HS students, the rate was 4.84%(95% *CI*: 4.04%-5.64%) 、8.34%(95% *CI*: 5.57%-11.12%) and 2.58%(95% *CI*: 0.98%-4.18%) during 2019-2023, respectively. (Table 3)

**Table 3.**
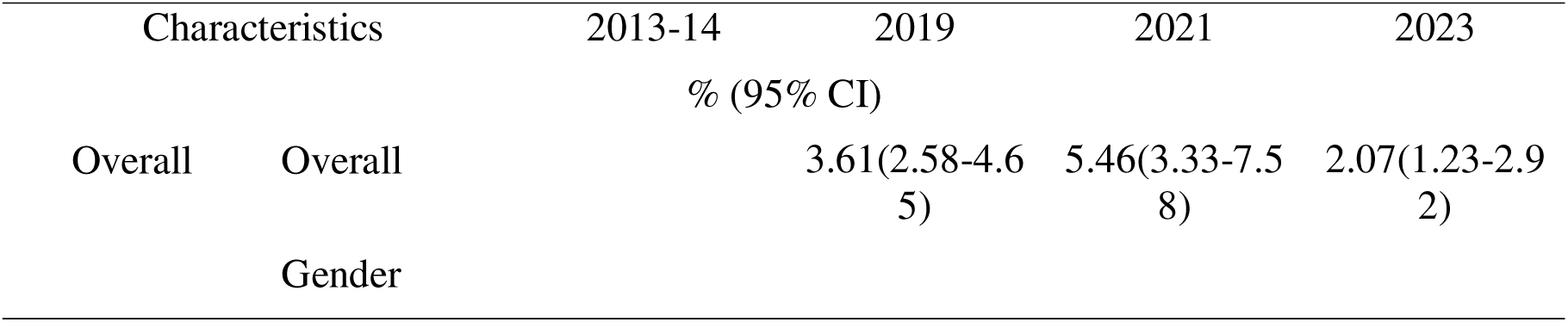
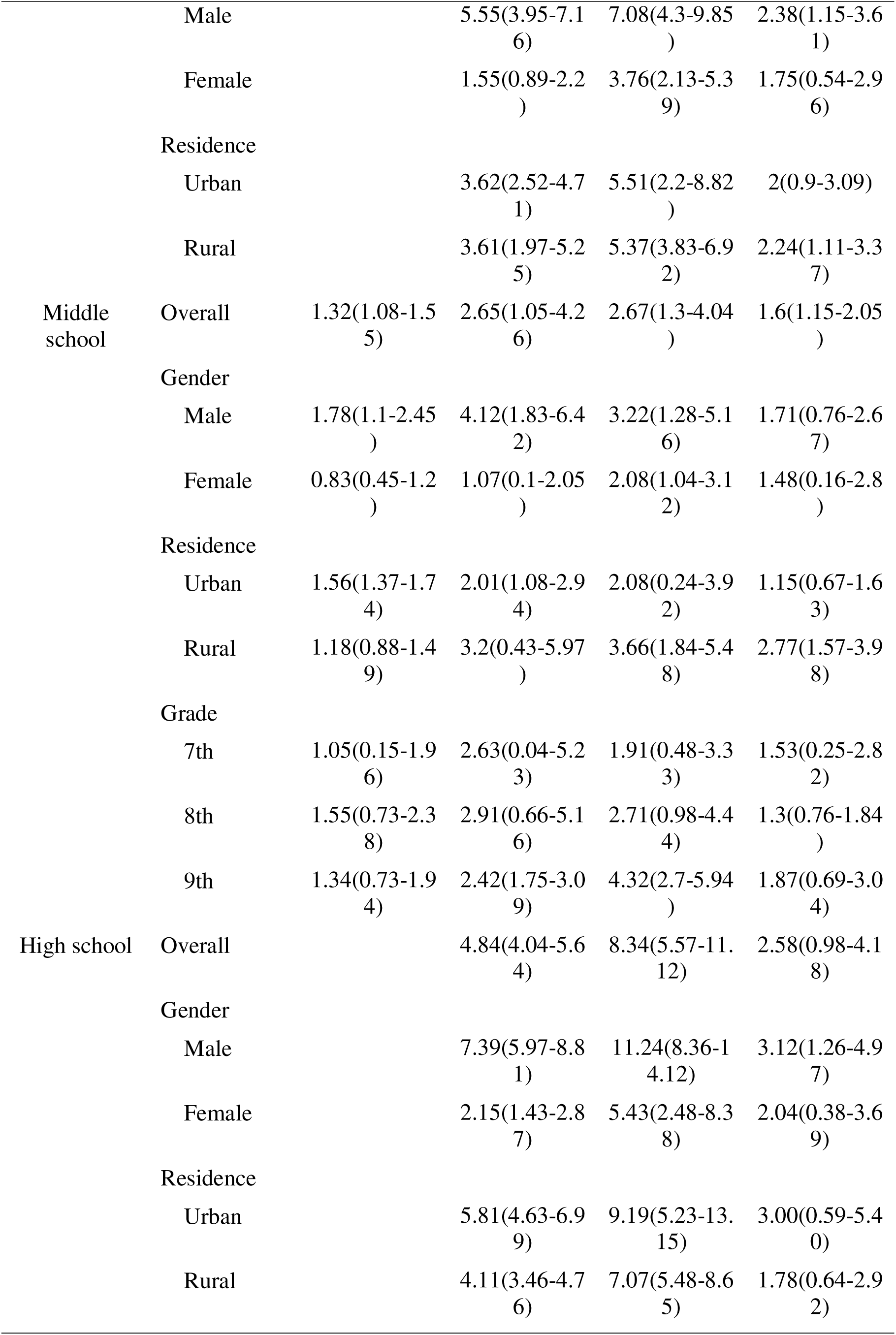
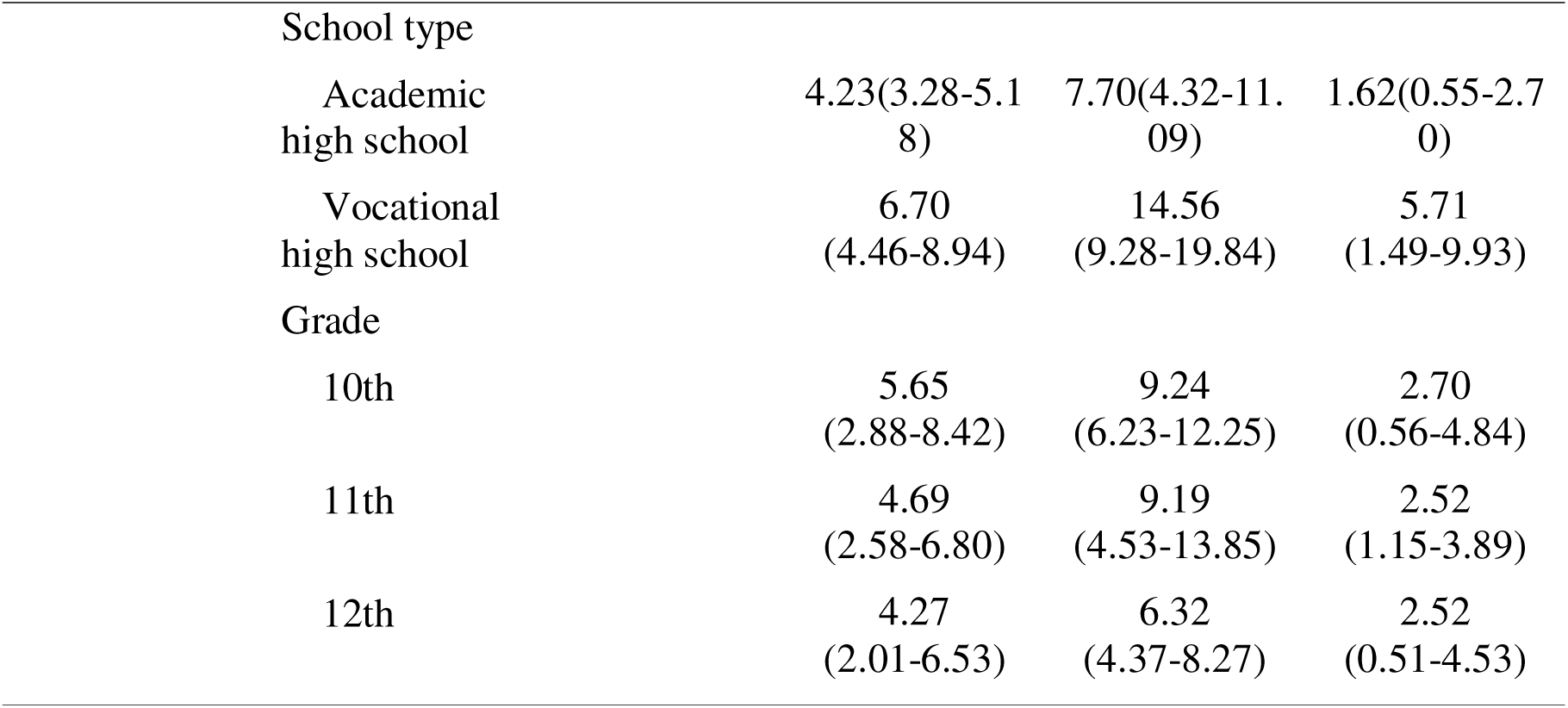
Current use of e-cigarettes among MS students and HS students in Heilongjiang, China.

### Current use of cigarette and/or e-cigarette among MS and HS students in Heilongjiang from 2014 to 2023

The current dual-use rate of cigarette and e-cigarette (the use rate of both cigarette and e-cigarette) among MS and HS students in Heilongjiang was 1.74% (95%*CI*: 1.06%-2.42%), 2.83% (95%*CI*: 1.58%-4.08%) and 1.03% (95%*CI*: 0.33%-1.73%), respectively. Among MS students, the rate was 0.31% (95%*CI*: 0.03%-0.59%), 0.94% (95%*CI*: 0.36%-1.51%), 1% (95%*CI*: 0.42%-1.57%), 0.48% (95%*CI*: 0.12%-0.85%), respectively from 2014 to 2023. Among HS students, the rate was 2.77% (95%*CI*: 1.92%-3.62%), 4.74% (95%*CI*: 3.09%-6.38%), 1.62% (95%*CI*: 0.4%-2.85%), respectively from 2019 to 2023.

The current rate of either cigarette or e-cigarette use among MS and HS students in Heilongjiang was 8.7% (95%*CI*: 5.58%-11.82%), 8.1% (95%*CI*: 5.02%-11.17%) and 5.76% (95%*CI*: 3.76%-7.76%), respectively from 2019 to 2023. Among MS students, the rate was 5.25% (95%*CI*: 2.84%-7.67%), 5.12% (95%*CI*: 2.37%-7.87%), 3.74% (95%*CI*: 1.88%-5.61%), 2.73% (95%*CI*: 1.86%-3.61%), respectively from 2014 to 2023. Among HS students, the rate was 13.27% (95%*CI*: 9.48%-17.06%), 12.62% (95%*CI*: 8.62%-16.62%), 9.01% (95%*CI*: 6.09%-11.94%), respectively from 2019 to 2023. (Table 4)

**Table 4.**
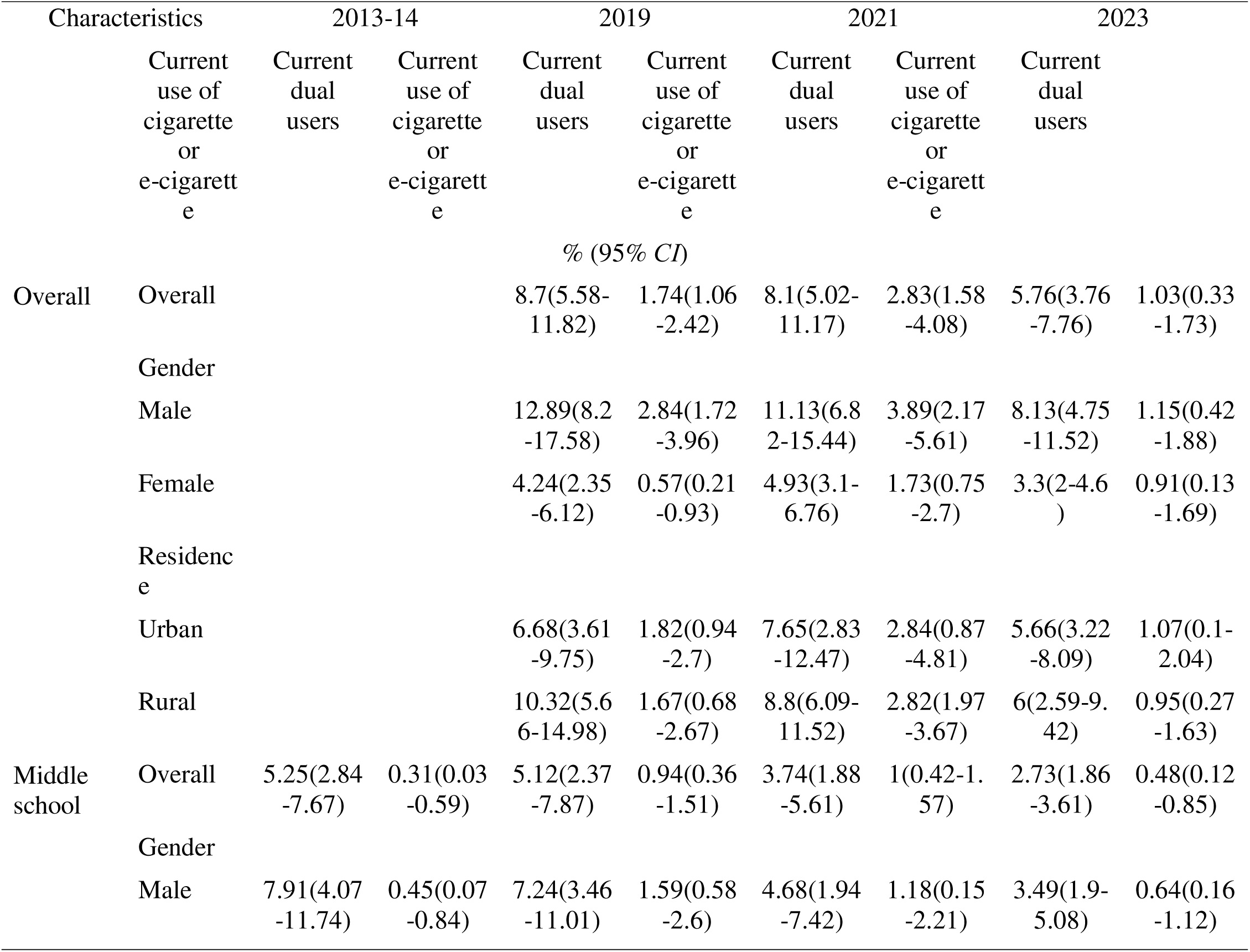
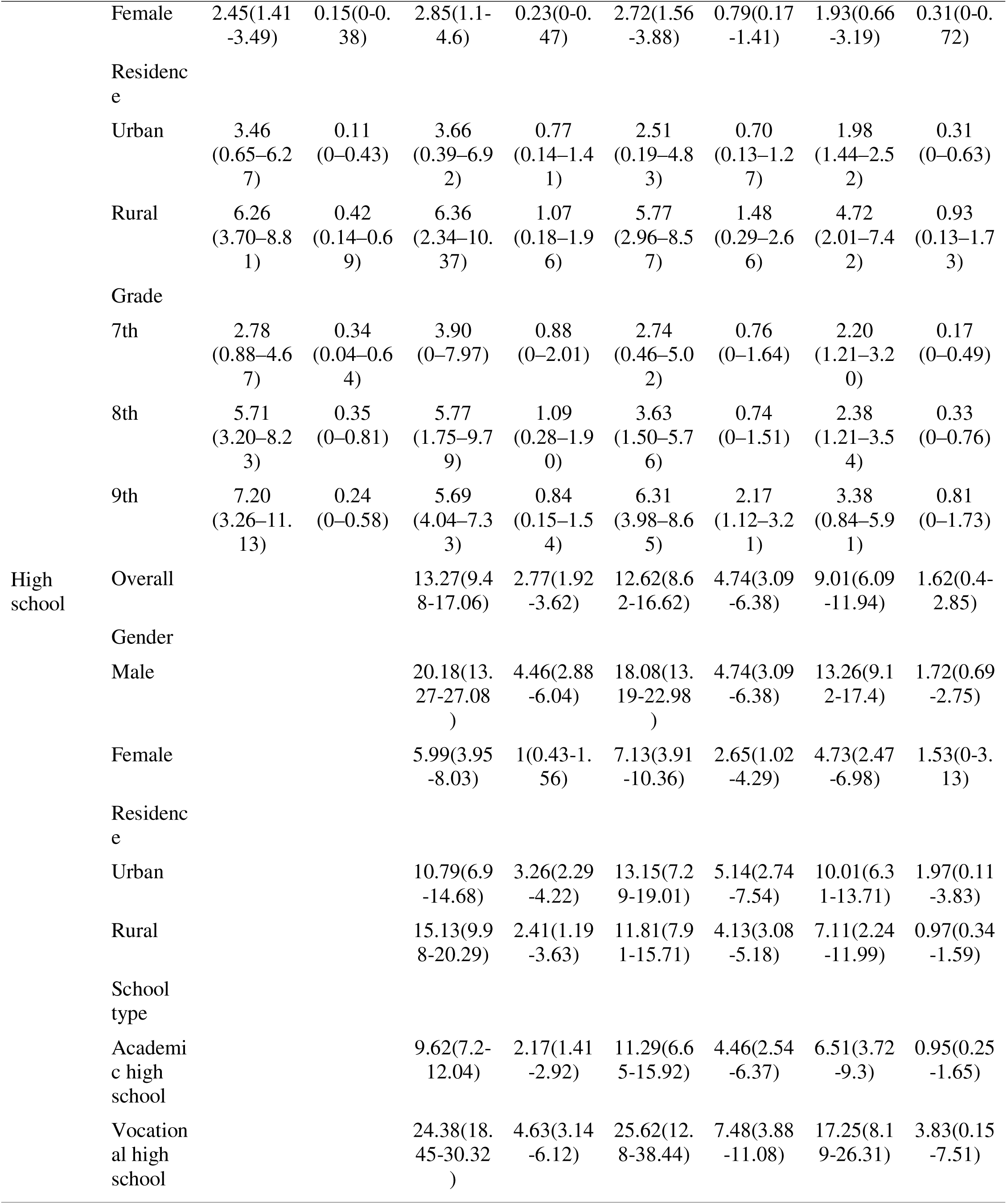
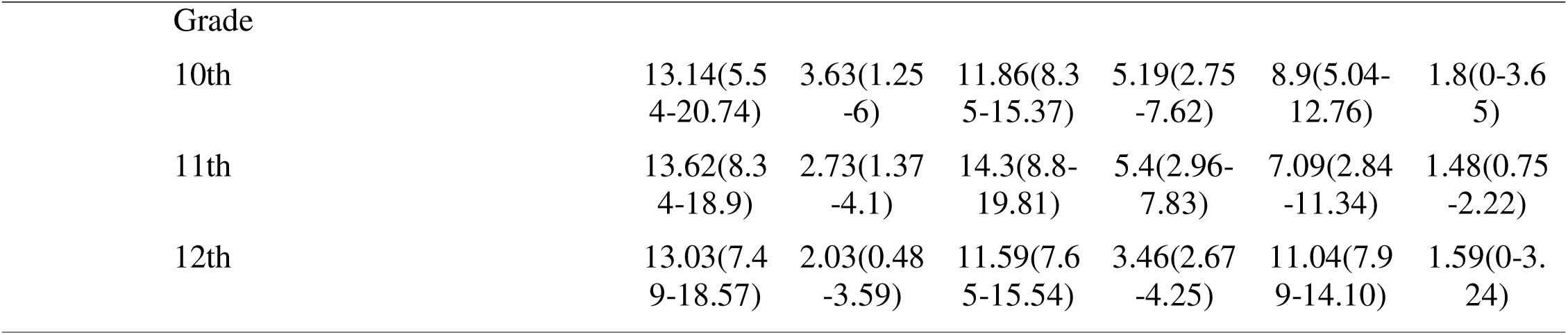
Current use of cigarette and/or e-cigarette among MS students and HS students in Heilongjiang, China.

### Association between e-cigarette and cigarette use among MS students and HS students in Heilongjiang Province during 2019-2023

From 2019 to 2023, among current cigarette smokers of MS and HS students in Heilongjiang Province, the proportion who had initially used e-cigarettes was 15.59% (*95% CI*: 11.78%-19.39%), 12.75% (*95% CI*: 4.50%-20.99%), and 21.80% (*95% CI*: 13.62%-29.98%), respectively. Among MS students, the proportion was 22.48% (*95% CI*: 9.44%-35.52%), 26.86% (*95% CI*: 6.15%-47.57%), and 21.50% (*95% CI*: 9.70%-33.30%), respectively. For HS students, the proportion was 12.97% (*95% CI*: 6.53%-19.40%), 9.85% (*95% CI*: 3.44%-16.27%), and 21.85% (*95% CI*: 12.09%-31.62%), respectively.

From 2019 to 2023, among cigarette and e-cigarette dual-users of MS and HS students in Heilongjiang Province, the proportion who had initiated tobacco use with e-cigarettes was 17.46% (*95% CI*: 8.61%-26.30%), 14.99% (*95% CI*: 3.93%-26.05%), and 28.41% (*95% CI*: 9.68%-47.13%), respectively. Among MS students, the proportion was 28.29% (*95% CI*: 12.40%-44.17%), 32.36% (*95% CI*: 10.04%-54.68%), and 22.89% (*95% CI*: 4.14%-41.63%), respectively. Among HS students, the proportion was 12.41% (*95% CI*: 2.76%-22.05%), 11.11% (*95% CI*: 3.26%-18.96%), and 30.37% (*95% CI*: 7.81%-52.93%), respectively. (Table 5)

**Table 5.**
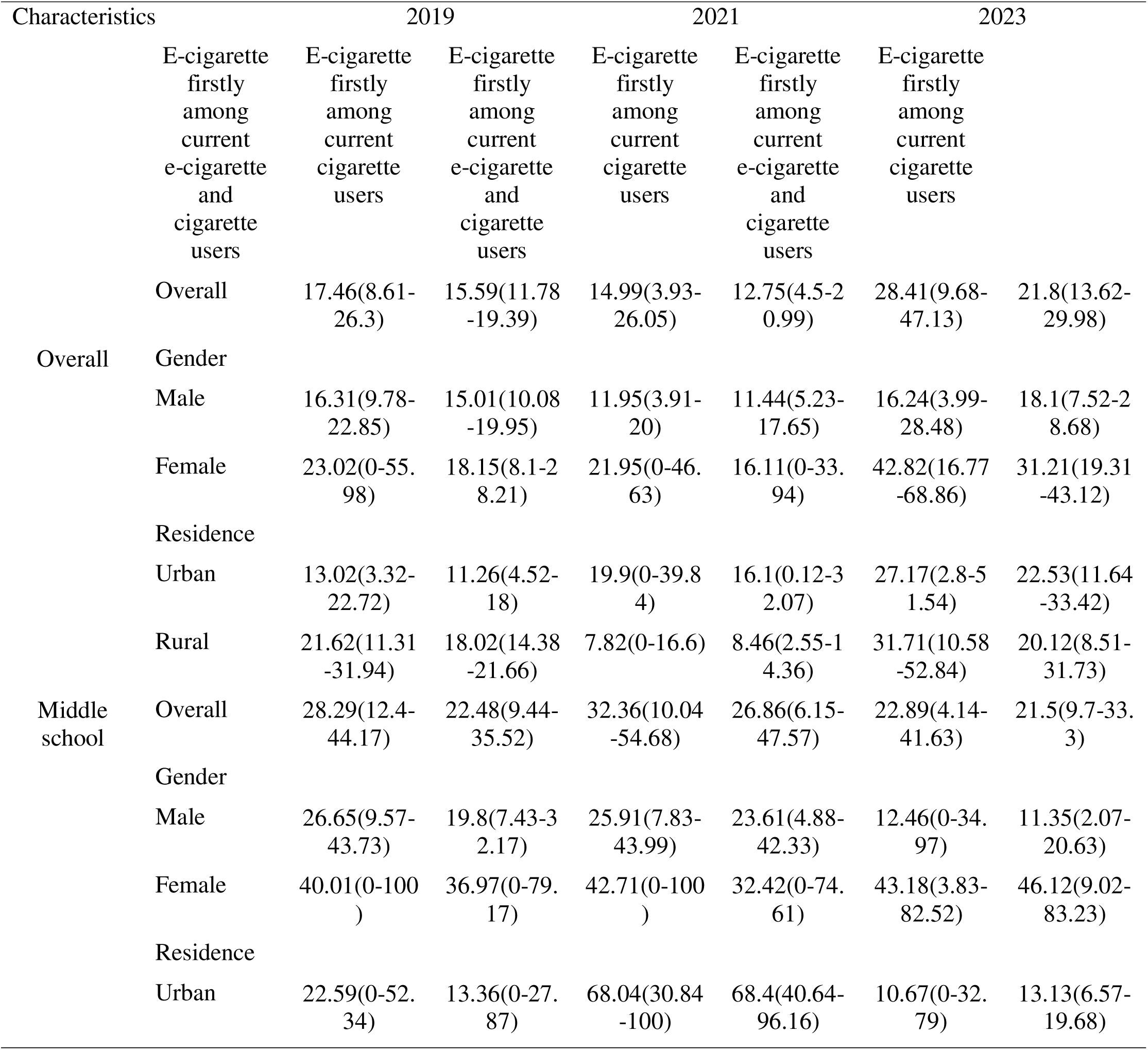
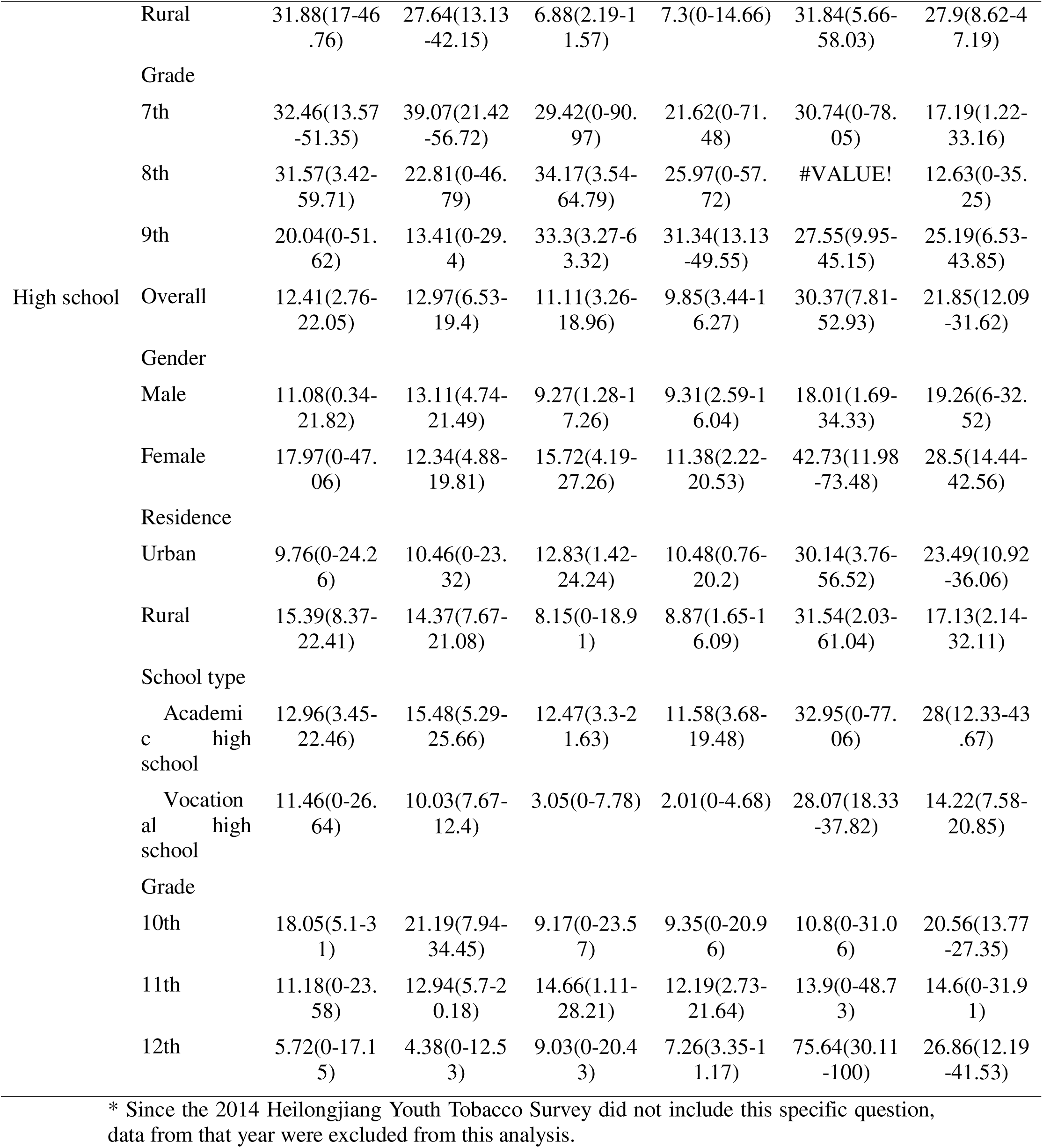
Association between e-cigarette and cigarette use among MS students and HS students in Heilongjiang, China.

### Analysis of factors associated with cigarette or e-cigarette use among MS and HS students in Heilongjiang, China

Univariate analysis of cigarette or e-cigarette use among MS and HS students in Heilongjiang, China in 2023 revealed several significant factors: gender, school type, weekly pocket money, parental smoking status, teachers’ smoking status, witnessing smoking on school premises, peers’ smoking status, awareness of secondhand smoke hazards, exposure to tobacco control information, exposure to tobacco advertisements, and exposure to smoking scenes in television/videos/movies (Table 6).

**Table 6.** Univariate analysis of current cigarette or e-cigarette use among middle school and high school students in Heilongjiang, China in 2023.

| Characteristics | Current use of cigarette or | $\chi^2$ | *P* |
| --- | --- | --- | --- |

|  | e-cigarette |  |  |
| --- | --- | --- | --- |
|  | (% , 95%CI) |  |  |
| Overall | 5.76(3.76-7.76) |  |  |
| Gender |  | 76.8411 | <0.0001 |
| Male | 8.13(4.75 ~ 11.52) |  |  |
| Female | 3.30(2.00 ~ 4.60) |  |  |
| Residence |  | 0.3409 | 0.8455 |
| Urban | 5.66(3.22 ~ 8.09) |  |  |
| Rural | 6.00(2.59 ~ 9.42) |  |  |
| School type |  | 260.006 | <0.0001 |
| Middle School | 2.73(1.86 ~ 3.61) |  |  |
| Academic high school | 6.51(3.72 ~ 9.30) |  |  |
| Vocational high school | 17.25(8.19 ~ 26.31) |  |  |
| Weekly pocket money (RMB) |  | 130.5158 | <0.0001 |
| 0 | 3.89 ( 1.72 ~ 6.05 ) |  |  |
| ≤10 (Not including 0) | 3.38 ( 1.45 ~ 5.32 ) |  |  |
| 11 ~ 30 | 4.35 ( 2.69 ~ 6.01 ) |  |  |
| 31 ~ 50 | 5.74 ( 3.67 ~ 7.80 ) |  |  |
| >50 | 11.46 ( 8.23 ~ 14.70 ) |  |  |
| Parents' smoking status |  | 100.8349 | <0.0001 |
| Neither parent smokes | 3.30 ( 1.71 ~ 4.90 ) |  |  |
| One parent smokes | 7.38 ( 5.78 ~ 8.99 ) |  |  |
| Both parents smoke | 12.99 ( 7.21 ~ 18.77 ) |  |  |
| Unknown | 8.36 ( 0.69 ~ 16.03 ) |  |  |
| Teachers' smoking |  | 235.9676 | <0.0001 |
| status |  |  |  |
| Almost daily | 15.70 ( 5.70 ~ 25.73 ) |  |  |
| Sometimes | 12.05 ( 7.51 ~ 16.58 ) |  |  |
| Never | 2.69 ( 1.82 ~ 3.56 ) |  |  |
| Unknown | 5.90 ( 4.02 ~ 7.78 ) |  |  |
| Observed someone<br>smoke on campus |  | 236.5651 | <0.0001 |
| Yes | 12.21 ( 8.10 ~ 16.33 ) |  |  |
| No | 2.97 ( 1.87 ~ 4.07 ) |  |  |
| Closest friends<br>smoke |  | 1385.274 | <0.0001 |
| None | 0.90 ( 0.22 ~ 1.57 ) |  |  |
| Some | 11.12 ( 7.88 ~ 14.36 ) |  |  |
| Most | 45.26 ( 38.63 ~ 51.89 ) |  |  |
| All | 45.58 ( 17.03 ~ 74.13 ) |  |  |
| Awareness of<br>secondhand smoking<br>hazards |  | 16.717 | 0.0009 |
| Yes | 4.91 ( 3.29 ~ 6.54 ) |  |  |
| No | 7.26 ( 4.24 ~ 10.28 ) |  |  |
| Exposed to<br>anti-tobacco<br>messages |  | 8.0199 | 0.0353 |
| Yes | 5.12 ( 3.76 ~ 6.49 ) |  |  |
| No | 6.73 ( 3.51 ~ 9.94 ) |  |  |
| Exposed to results of<br>tobacco use in<br>academic settings |  | 6.1195 | 0.3477 |
| Yes | 5.32 ( 3.22 ~ 7.41 ) |  |  |
| No | 6.75 ( 4.78 ~ 8.73 ) |  |  |
| Unknown | 5.26 ( 1.77 ~ 8.75 ) |  |  |
| Exposed to tobacco advertisement |  | 134.9397 | <0.0001 |
| Yes | 4.31 ( 2.59 ~ 6.03 ) |  |  |
| No | 3.16 ( 0.59 ~ 5.73 ) |  |  |
| Not applicable | 11.55 ( 8.54 ~ 14.57 ) |  |  |
| Exposed to smoking scenes in TV/videos/movies |  | 98.3775 | <0.0001 |
| Yes | 2.81 ( 1.34 ~ 4.29 ) |  |  |
| No | 4.21 ( 2.16 ~ 6.25 ) |  |  |
| Not applicable | 8.94 ( 6.37 ~ 11.50 ) |  |  |

Multivariable logistic regression analysis was performed with exclusive cigarette or e-cigarette use among MS and HS students as the dependent variable (yes=1, no=0), incorporating all significant predictors identified in the univariate analysis. The results demonstrated that males had 1.815 times higher odds of use than females (95% *CI*: 1.090–3.021). Compared with MS students, AHS students and VHS students exhibited 1.613-fold (95% *CI*: 1.129–2.305) and 3.195-fold (95% *CI*: 1.434–7.121) higher odds of use, respectively. Students reporting >50 Yuan weekly pocket money had 1.564 times greater odds than those with no pocket money (95% *CI*: 1.240–1.972). Those with one or both parents smoking showed 1.423-fold (95% CI: 1.154–1.755) and 2.874-fold (95% CI: 2.112–3.911) increased odds versus students with non-smoking parents. Students witnessing smoking on school premises had 1.503 times higher odds than those not exposed (95% *CI*: 1.276–1.770). Compared to students with no smoking friends, those reporting some, most, or all friends smoking demonstrated 6.868-fold (95% *CI*: 4.046–11.657), 29.23-fold (95% *CI*: 14.726–58.018), and 38.761-fold (95% *CI*: 17.872–84.069) increased odds, respectively.

Students with incorrect knowledge of secondhand smoke hazards had 1.838 times higher odds than those with correct knowledge (95% *CI*: 1.468–2.303). All associations were statistically significant (*P* < 0.05) (Table 7).

**Table 7.**
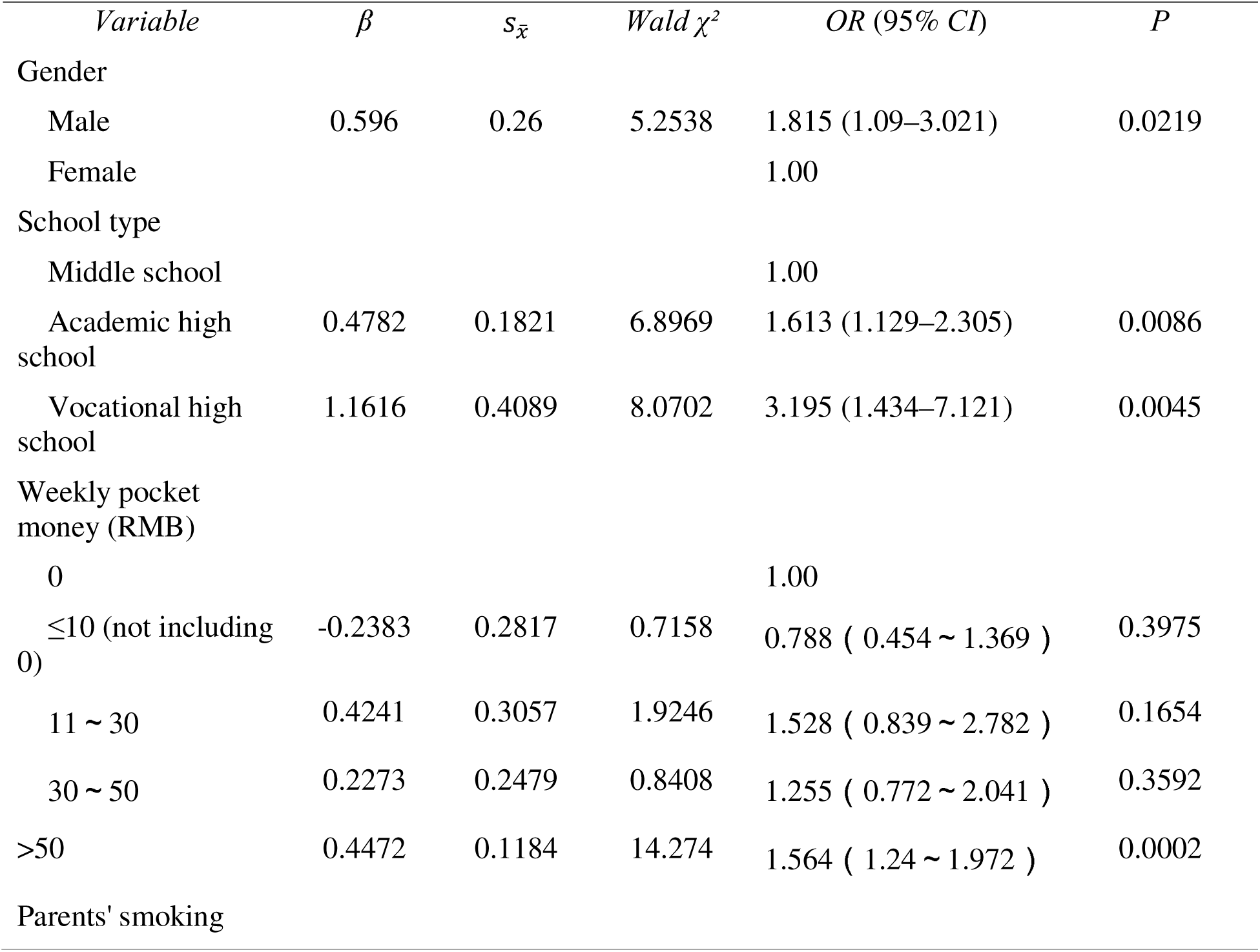
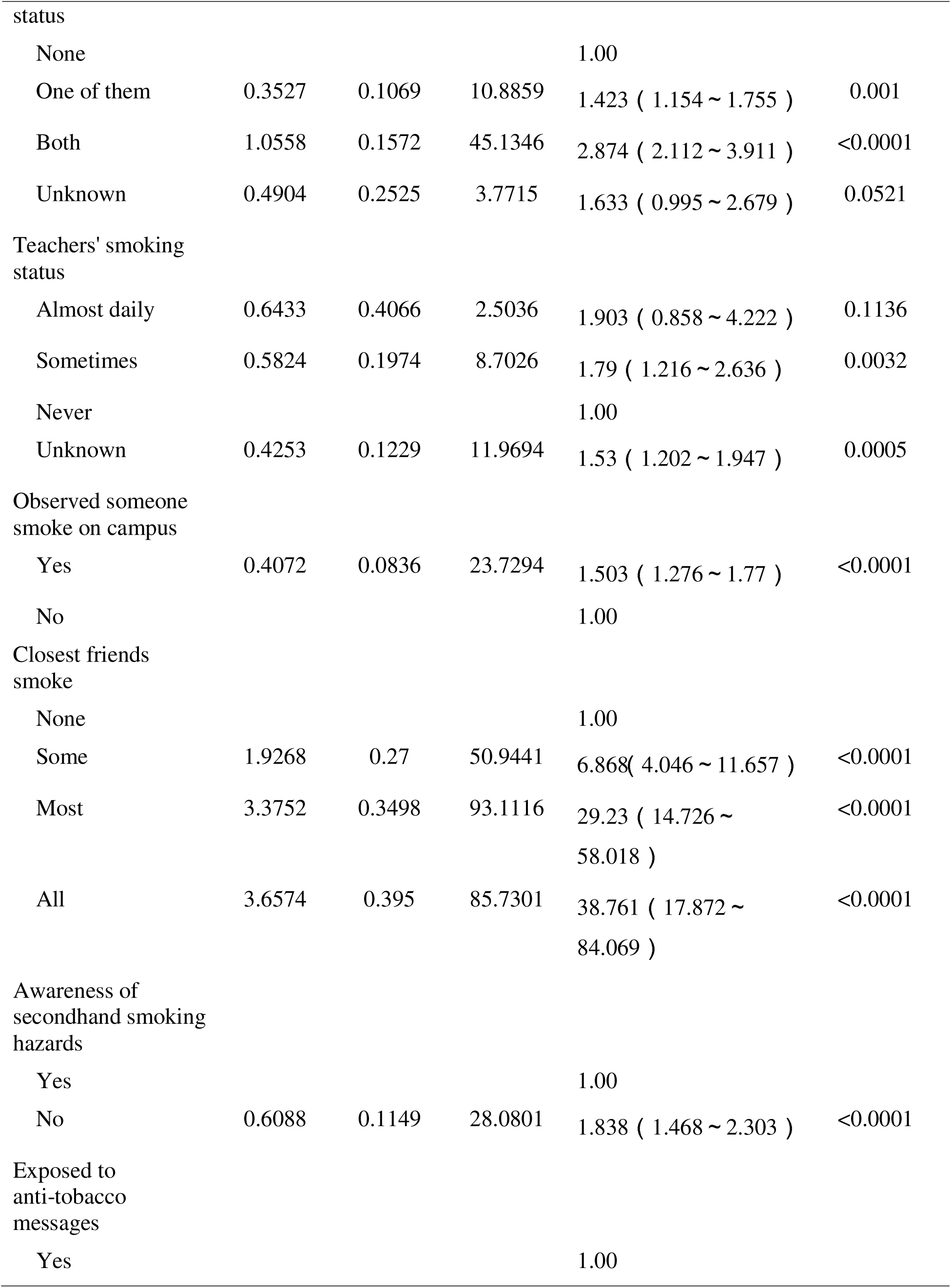
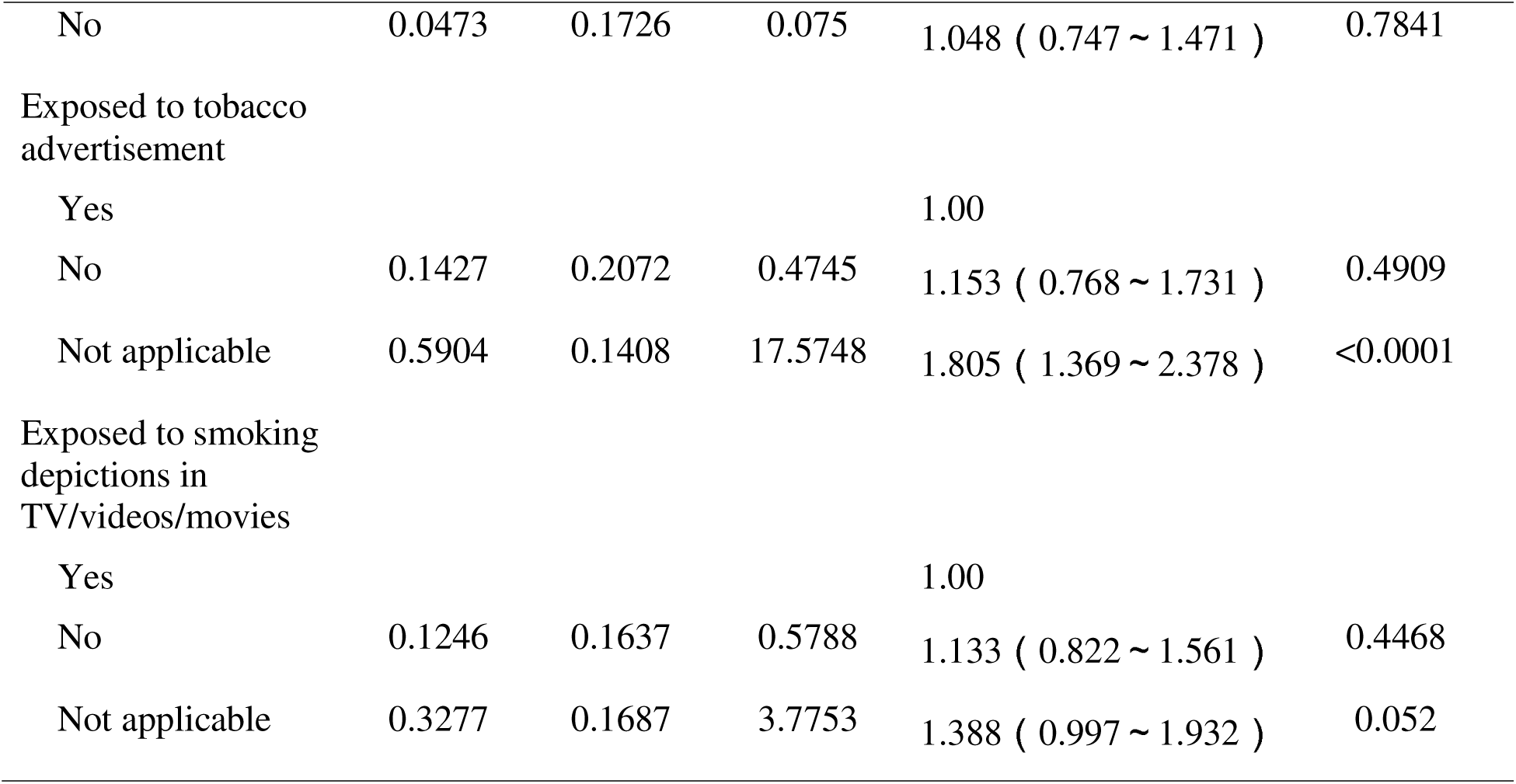
Multivariate analysis of cigarette and e-cigarette use among MS students and HS students in Heilongjiang, China in 2023.

## DISCUSSION

Based on the youth tobacco survey results from Heilongjiang Province (2014-2023), current smoking rates among MS and HS students demonstrated a significant downward trajectory. The aggregate current smoking prevalence across both MS and HS populations decreased substantially from 6.79% in 2019 to 4.70% in 2023. When stratified by educational level, MS students exhibited a pronounced decline from 4.22% in 2014 to 1.6% in 2023, while HS students showed a parallel reduction from 11.19% in 2019 to 8.02% in 2023. These declining prevalence patterns align with trends documented in the National Youth Tobacco Survey (NYTS) ^12^. Such favorable trajectories provide compelling evidence that the “Tobacco-Free Campus” policy implementation has yielded preliminary success in curtailing cigarette consumption among adolescent students in Heilongjiang Province. The policy foundation for these initiatives was established in 2010 when the General Office of the Ministry of Education and the General Office of the Ministry of Health jointly issued the “Opinions on Further Strengthening Tobacco Control in Schools” ^21^, which mandated comprehensive indoor and campus-wide smoking prohibitions across secondary vocational schools, primary/secondary schools, and kindergartens—formally inaugurating the “Tobacco-Free School” initiative.

Subsequently, in 2014, the former Ministry of Education reinforced these measures through the “Notice on Matters Related to Smoking Bans in All Types of Schools Nationwide” ^22^, which emphasized the importance of regionally tailored implementation of Tobacco-Free School programs.

In 2021, the Office of the National Patriotic Health Campaign Committee and the Office of the Healthy China Action Promotion Committee jointly issued the *Specifications for Constructing Healthy Villages (Healthy Cells) and Healthy Towns/Counties (Trial)*. Schools were designated as a critical component of “Healthy Cell” development, with Article 21 of the “Healthy School” construction specifications explicitly mandating: “Establish tobacco-free schools; display prominent no-smoking signs at entrances and key campus areas; ensure no smoking on campus and prohibit tobacco advertising, promotions, or sponsorships. Teachers and students should lead by example in complying with tobacco control regulations in public spaces. Tobacco and alcohol sales to minors by businesses near campuses are prohibited“^23^. These regulatory frameworks have provided substantive policy support for reducing youth smoking prevalence. Notably, from 2019 to 2023, current smoking rates among high school students consistently exceeded those of middle school students in Heilongjiang Province.

Compared to their middle school counterparts, high school students demonstrated greater psychological maturity and access to diverse information channels. Moreover, the intensified academic pressure and associated psychological distress experienced by high school students likely functioned as significant contributing factors to smoking initiation. Addressing this psychological-to-physiological pathway necessitates a comprehensive five-party intervention strategy integrating school personnel, administrators, parents, community stakeholders, and broader societal entities. This multifaceted approach should prioritize evidence-based stress management counseling for high school students while emphatically countering the misconception that tobacco use alleviates psychological distress.

From 2019 to 2023, e-cigarette use among middle and high school students in Heilongjiang Province demonstrated an initial increase followed by a subsequent decline, a pattern observed consistently across both student populations. Compared to 2019 baseline levels, current e-cigarette use rates among both middle and high school students increased significantly by 2021, then decreased substantially by 2023. This trajectory suggests that early tobacco control strategies in Heilongjiang primarily targeted conventional cigarettes while failing to adequately address e-cigarette regulation.

Consequently, current e-cigarette use rates among students increased markedly between 2019 and 2021. The psychological “mere-exposure effect” may explain this phenomenon, whereby repeated exposure to stimuli generates positive evaluations or preferences ^24^. Students’ exposure to e-cigarette use or advertisements across multiple settings and channels likely reinforced positive impressions toward these products. Curiosity-driven experimentation may have followed, with some students progressing to regular use. Given youth populations’ greater receptiveness to novel products compared to adults, e-cigarette adoption rates were notably elevated in this demographic. Educational institutions, parents, and society gradually recognized the health risks e-cigarettes posed to youth and their long-term implications. In 2019, eight national agencies including the National Health Commission and the Publicity Department of the CPC Central Committee jointly issued the Notice on Further Strengthening Adolescent Tobacco Control ^25^. That same year, the State Tobacco Monopoly Administration and the State Administration for Market Regulation released the Notice on Further Protecting Minors from E-cigarette Hazards ^26^, bringing youth e-cigarette use under regulatory oversight and advocating for youth avoidance of these products. Nevertheless, e-cigarette popularity surged among youth during 2019–2021, with the increasing trend among Heilongjiang students driven by multiple factors: extensive promotion through new media channels, accessible online and offline purchasing platforms, and policy implementation delays following the emergence of novel products. A regulatory turning point occurred in November 2021 when the State Council promulgated the Decision on Amending the Implementation Regulations of the Tobacco Monopoly Law of the People’s Republic of China. This revision explicitly stipulated that “new tobacco products such as e-cigarettes shall be subject to the relevant provisions of these Regulations concerning cigarettes“^27^. Subsequently, the mandatory E-cigarette National Standard (GB 41700-2022) took effect on October 1, 2022. As China’s first compulsory national standard for e-cigarettes, it aimed to reduce product appeal while enhancing protections for minors, establishing a new regulatory baseline ^28^ and strengthening school-based e-cigarette control. Educational institutions progressively incorporated e-cigarettes into existing “Tobacco-Free Campus” frameworks, implementing equivalent control measures for both e-cigarettes and conventional cigarettes. Daily tobacco control education intensified messaging about e-cigarette risks, transforming student attitudes and behaviors—shifting perceptions from e-cigarette use as fashionable to tobacco rejection as the aspirational norm. Concurrently, market regulatory authorities standardized and supervised e-cigarette sales channels, while educational and health departments maintained heightened surveillance of campus e-cigarette trends. This multi-departmental regulatory approach and cross-sectoral collaboration drove the significant decline in current e-cigarette use rates among middle and high school students during 2021–2023.

From 2014 to 2023, dual-use of conventional cigarettes and e-cigarettes among students in Heilongjiang Province exhibited a pattern of initial increase followed by subsequent decline, mirroring provincial e-cigarette usage trends. This pattern was consistent across both MS and HS student populations. Concurrently, exclusive use of either cigarettes or e-cigarettes decreased across all student demographics, suggesting e-cigarettes’ profound influence on youth tobacco consumption patterns. Notably, e-cigarette usage demonstrated a stronger impact than conventional cigarette use on dual-use rates, highlighting the critical importance of policies restricting youth access to e-cigarettes ^29^. A comprehensive regulatory framework has emerged, including the Law of the People’s Republic of China on the Protection of Minors, which stipulates that “no tobacco-branded products may be sold to minors” ^30^, the revised Implementation Regulations of the Tobacco Monopoly Law of the People’s Republic of China ^27^, the E-cigarette Management Measures ^31^, the mandatory E-cigarette National Standard (GB 41700-2022) ^28^, and supplementary policies—collectively forming a comprehensive “1+2+N” e-cigarette regulatory system ^32^. This integrated approach specifically targets youth protection from e-cigarette hazards. Effective safeguarding of youth physical and mental health necessitates multi-policy synergy regulating the entire e-cigarette lifecycle: from production and distribution to marketing channels and promotional activities ^33^. E-cigarettes represent a novel nicotine delivery mechanism—not merely cigarette substitutes as commonly perceived—yet both globally and domestically, nicotine has increasingly been introduced to youth populations through these devices ^34^. Research indicates that e-cigarettes may function as catalysts for conventional cigarette initiation among youth while potentially fostering group-based usage patterns and amplifying social influence within school environments ^35^.

Educational institutions should strategically leverage the substantial social capital of student leaders to redirect peer influence positively, transforming these dynamics into advocacy tools that redefine campus norms. Furthermore, dynamic updates to school tobacco control strategies could facilitate youth progression from passive information reception to belief formation and ultimately behavioral change. Continuous reinforcement of tobacco-related health hazards and cessation challenges through sustained messaging would strengthen risk perception and solidify commitment to tobacco avoidance among youth populations ^36^.

From 2019 to 2023, the proportion of current cigarette users and dual-users who initiated tobacco use with e-cigarettes first decreased then increased among middle and high school students in Heilongjiang. According to 2023 survey data, the rate of e-cigarette initiation among high school students (21.85%) exceeded that of middle school students (21.50%) among current cigarette users. Similarly, among current dual-users, more high school students (30.37%) initiated with e-cigarettes compared to middle school students (22.89%). This pattern suggests that without e-cigarette availability, some youth might never have initiated tobacco use. Following their introduction in Western markets in 2006, e-cigarettes rapidly gained popularity as novel nicotine alternatives ^37^. Existing research conclusively demonstrates that youth—whether exclusive e-cigarette users, exclusive cigarette smokers, or dual-users—face greater exposure to toxins and carcinogens than non-users ^7, 38^. Moreover, e-cigarette use may increase smoking susceptibility or prompt cigarette initiation among youth ^17, 39–41^. Consequently, early school-based interventions are paramount. Beyond maintaining tobacco-free campus initiatives and updating control strategies, cross-departmental collaboration between national health and education authorities is essential. Key priorities include: integrating comprehensive tobacco risk education—including e-cigarette-specific content—into mandatory junior and senior secondary health curricula, developing unified national teaching materials, lesson plans, and instructional hours, establishing student-centered knowledge dissemination networks that extend outreach to families, peers, communities, and workplaces, leveraging these networks to create multiplier effects through interpersonal communication.

Analysis of factors influencing cigarette or e-cigarette use among middle and high school students in Heilongjiang Province, based exclusively on 2023 survey data, identified several significant determinants: gender, school type, weekly disposable pocket money, parental smoking status (single or dual-parent), observed smoking on campus, peer smoking behavior, and accurate perception of secondhand smoke hazards. These findings underscore the substantial influence of one’s immediate social circle during this psychologically sensitive developmental period. Existing research indicates that cohabitating with smokers significantly elevates risks of both tobacco initiation and sustained use. Parental or peer smoking behaviors not only facilitate smoking initiation but also increase consumption among current users, with risks escalating proportionally to exposure duration ^42^. The observed impacts of household weekly disposable pocket money and parental smoking status further confirm that effective youth tobacco control interventions fundamentally require family engagement as a critical component in youth development. The Planning and Development Department of Informatization formally endorsed this approach through its Notice on Promoting Tobacco-Free Household Initiatives, explicitly stating: “Tobacco-free households constitute the foundation of a smoke-free society. Establishing such environments is paramount for cultivating healthier social norms and safeguarding family health” ^43^. As an essential element of “Healthy Cell” initiatives, incorporating tobacco-free households into governmental planning frameworks promises to foster smoke-free domestic environments while strengthening youth tobacco control awareness.

Future school-based interventions should consequently prioritize integrating tobacco control with psychological education to address three developmental transitions: from initial awareness formation to conviction strengthening, followed by enhanced behavioral discernment. This approach enabled students to resist negative peer influences while developing resilience against detrimental social behaviors. Furthermore, cultivating diplomatic smoking intervention skills—particularly in indoor public spaces—represented a critical emotional intelligence competency for contemporary youth. Educators should systematically empower students to confidently articulate “Please refrain from smoking” in public settings, thereby developing youth capacity for responsible intervention. This competence derived from three foundational elements: comprehensive tobacco hazard knowledge, advanced emotional intelligence training, and conscientious prioritization of communal health—all of these contents required systematic knowledge dissemination within educational institutions.

**Accurate understanding of secondhand smoke hazards emerged as a particularly significant factor influencing exclusive cigarette/e-cigarette use.** Empirical evidence confirms a positive correlation between misconceptions about cigarette/e-cigarette hazards and youth experimentation ^44^, while correct hazard knowledge demonstrates a negative correlation. This necessitates refined risk communication strategies in future campaigns: tobacco risk education should deconstruct e-cigarette dangers through targeted messaging about health impacts of aerosol constituents (e.g., ultrafine particles, carbonyl compounds) and pathophysiological effects of e-liquid components (propylene glycol, nicotine salts) on specific organ systems ^45^. Such granular knowledge dissemination would overcome limitations of traditional awareness campaigns, enable deep knowledge penetration, and ultimately foster interpersonal diffusion of tobacco risk information.

## Limitations

The questionnaires were self-administered by MS students and HS students, which cannot guarantee the accuracy of their subjective interpretation during completion. This approach may introduce underreporting of tobacco use behaviors and recall bias to some extent. Furthermore, the study exclusively involved school-attending MS students and HS students, excluding non-school-attending youth within the same age demographic. This sampling limitation may introduce selection bias, potentially resulting in tobacco control policies and strategies that do not fully address this specific population. As a cross-sectional design, this study cannot establish causal inference. The temporal sequence of cigarette and e-cigarette initiation cannot be determined, precluding investigation into potential causal pathways between these behaviors.

## Conclusions

Current cigarette use among middle and high school students in Heilongjiang Province declined from 2014 to 2023, while e-cigarette use demonstrated an initial increase through 2021 before subsequently decreasing. These trends indicate that tobacco-free campus policies and e-cigarette regulations are beginning to yield measurable results. However, the complex patterns observed in dual-use behaviors and product-switching sequences underscore the need for comprehensive, multi-faceted interventions. Future tobacco control efforts must prioritize enhanced cross-departmental collaboration, innovative knowledge dissemination strategies tailored to provincial contexts, and strengthened enforcement mechanisms. These evidence-based approaches will effectively advance the establishment of comprehensive tobacco-free campuses, tobacco-free households, and ultimately contribute to achieving a tobacco-free society.

## ETHICAL APPROVAL AND INFORMED CONSENT

The 2019 China National Youth Tobacco Survey received approval from the Chinese Center for Disease Control and Prevention Institutional Review Board (Approval Number: 202008; Date: May 26, 2020). The 2021 China National Youth Tobacco Survey was approved by the Chinese Center for Disease Control and Prevention Institutional Review Board (Approval Number: 202110; Date: March 25, 2021). The 2023 China National Youth Tobacco Survey obtained approval from the Chinese Center for Disease Control and Prevention Institutional Review Board (Approval Number: 202301; Date: March 6, 2023). Survey questionnaire completion constituted informed consent. All methodological procedures were conducted in accordance with relevant guidelines and regulations.

## DATA AVAILABILITY

The datasets supporting this research cannot be made publicly available due to privacy restrictions and other confidentiality considerations.

## AUTHORS’ CONTRIBUTIONS

Linlin Jiang and Ying Wang conducted data cleaning, performed statistical analyses, and drafted the initial manuscript. Xinbo Di designed the study methodology, supervised manuscript preparation, and conducted comprehensive revisions. All authors contributed substantially to data interpretation and preparation of the final manuscript version.

## PROVENANCE AND PEER REVIEW

The authors did not commission external peer review.

